# Characterization and Profiling of Gut Bacterial Microbiome and Pathobionts among HIV-negative and HIV-infected individuals in Cameroon

**DOI:** 10.1101/2022.05.24.22275521

**Authors:** Simon Eyongabane Ako, Celine N. Nkenfou, Jules N. Assob, Thumamo Benjamin Pokam, Christopher Njopin, Enoh Jude Eteneneng, Cho Frederick Nchang, Mbanya Gladice Mbanya, Woguia Gilles-Fils, Ngoume Moukoma Y. Franck, Eric Achidi Akum

## Abstract

**Background:** Knowledge of the core gut microbiome among Cameroonians is a preliminary step for a better implementation of treatment strategies to correct dysbiosis and improve health care management. HIV infection has continued to cause high mortality among those infected, but the types and frequency of human gut microbiota associated with or without HIV/AIDS presence have not been identified in the adult populations in Cameroon.

**Methods:** This was a case-control and comparative study design that ran from June 2018 to September 2019. Stool Samples were purposively collected from 40 participants (15 HIV-negative and 25 HIV-positive) for the 16S rRNA gene sequencing on the next-generation Illumina® MiSeq™ sequencer. Blood samples were collected for HIV determine testing, CD4 Tcell count and HIV viral load. Sequences were clustered into operational taxonomic units (OTUs) at ninety-nine percent identity and their representatives were accustomed to using a phylogenetic approach.

**Results:** The study showed a phylogenetic taxonomy of the gut microbiome communities in two kingdoms (Archea and Bacteria) and eight Phylum [Firmicutes (44.7%), Bacteroidetes (43.7%), Proteobacteria (8.7%), Actinobacteria (1%), Fusobacteria (0.2%), Euryarchaeota (0.01%), Synergistetes (0.01%), Verrucomicrobia (0.01%) and unclassified phylum (1.7%)]. A total of 347 gut microbiota species were identified, 55 unique species/oligotypes and 28 pathobionts from the study population. HIV infection was characterized by significant gut microbiota compositional changes with HIV-positive less diverse (56 strains absent) with significant increased OTUs of the butyrate-producing microbiome species as compared to HIV-negative individuals (p=0.001).

**Conclusions:** A profile of 347 gut microbiome bacterial species where identified in the Cameroonian community. Particularly, from the 347 gut microbiome bacterial species profiled, eight phyla were identified, with 55 unique species/oligotypes containing more than one sequence and 28 pathobionts. A host of unknown/unclassified gut microbiome bacterial species were also noted circulating among the study population.

**Key Points:** There is high diversity and specie uniqueness of the gut microbiome in Cameroon. Functionality of the gut microbiome needs to be investigated.

There are gut pathobionts circulating among HIV-infected and HIV-negative individuals in Cameroon. The origin of pathobionts is a call for concern.

Unique gut microbiome OTU sequences are significantly high among HIV-infected. Emerging strains of new microorganisms are on a rise.

## BACKGROUND

The Gut microbiota comprises a consortium of microorganisms that are viewed as permanent occupants of the human intestinal tract. These microorganisms incorporate bacteria, microbial eukaryotes, viruses/phages, and archaea, together with their genes are typically described as microbiome [1]. Various components add to the foundation of the human gut microbiota during development and of which, diet is considered one of the fundamental determinants of informing the gut microbiota over the growth time. Change in gut microbiota composition (dysbiosis) has been related to the pathogenesis of numerous inflammatory conditions, coronary illness, diabetes, and malignancy [2]. The life systems and physiology of the human gastrointestinal tract provide a broad region (250 — 400m^2^) for different host collaboration processes that control the integrity of the host. Given the immense nature of the gastrointestinal tract, the assessed gut microbiota population possessing the zone surpasses 10^14^ count. Summarily the gut comprises roughly multiple times more microbial cells than human cells, and multiple times the microbiome when compared with the human genome [3]. The past depiction used to construe this tremendous colonization rates of microorganisms in humans were described as “Superorganism”. Works carried out on a revised estimate for human and bacteria cells in 2016, revealed that the proportion of human to bacterial cells was 1:1[4]. With the expanding understanding of the gut microbiota’s role in maintaining health through major physiological capacities like modulating host immunity and safeguarding against remote substances and pathogens, more works are needed to elucidate the various gut microbiota present in different communities and their gut population level among the individuals in that community. A few natural elements have been associated with shaping the microbiota including topographical areas, living courses of action (urban or rural) [5], and HIV infection/treatment [6].In 2016, Cameroon had 32 000 (22 000–41 000) new HIV infections and 29 000 (25 000–33 000) AIDS-related deaths. There were 540 000 (470 000–650 000) people living with HIV in 2018 [7]. The gut microbiome for a Cameroonian population needs to be studied due to the predominance in an urban Cameroonian setting of, HIV-1 CRF02_AG viruses alongside viruses belonging to known HIV-1M clades, URFs, and currently unclassified divergent lineages [8]. Profiling the gut microbiome of HIV patients in this community will offer a database containing various gut microbiome species that are common among HIV patients.

Although Phyla Bacteroidetes and Proteobacteria have been shown to make up the majority of gut microbiota composition, a few species among them have been reported as pathobionts [9]. In developed countries, where most works on the human microbiome have been performed, findings have differently uncovered that HIV-infected people have a high abundance of Prevotella and fewer Bacteroides at the genus level than do uninfected controls [10]. Few studies have uncovered a similar loss of diversity-related to HIV infection status [11]. In any case, works on the relationship between HIV disease and diminished microbiota diversity have not been reliably observed in the adult Cameroonian population. A previous work, on capturing the gut microbiota dysbiotic pattern among HIV negative individuals, and HIV-positive patients with /or without first-line ARV and cotrimoxazole prophylaxis treatment through culture-dependent technique in an adult Cameroonian population, provided early evidence of dysbiosis in the gut flora [12]. The study had the limitation of characterizing all the genera present. Therefore, there is an absolute need to conduct additional studies that will identify and profile the gut microbiome in adult HIV/AIDS patients compared to HIV-negative individuals. The gut microbiome profile of HIV patients will provide baseline data wherein valuable information will be used to improve the management of HIV/AIDS patients. Understanding the dysbiosis in HIV patients will help enlighten health care providers on new measures for improving the management of HIV patients. Thus, advice on probiotics might be implemented based on the degree of dysbiosis observed following these and other studies.

## Material and Methods

### Study design

This was a case-control and comparative study including both HIV seropositive and seronegative participants. Totally, from a cohort of 320 volunteer adult participants (100 HIV-negative and 220 HIV-positive), 25 study participants were purposively recruited from the HIV treatment Unit (UPEC) at the Buea Regional Hospital, Southwest Region, Cameroon. Additionally, we included 15 HIV-negative controls (negative). Inclusion criteria were age >18 years, HIV positive for at least 6 months, and no ongoing HIV-related complications. Exclusion criteria were inflammatory bowel disease or infectious gastroenteritis within the last four weeks.

All study participants gave written informed consent. All the work and experiments were performed following relevant guidelines, and regulations and with the Declaration of Helsinki. The study was approved by the Institutional Review Board (IRB) of the Faculty of Health Sciences (FHS) of the University of Buea, Cameroon Ref N°: 2018/826-06/UB/SG/IRB/FHS.

### Stool sample collection

Each individual was requested for a stool sample. Study participants were given a sterile stool collection container with appropriate instructions on how to deposit the sample without any contamination from urine. The container was closed and brought out immediately (within 5 minutes) by the participants. The Stool samples were transported on an ice bath with an ice pack from the collection site (Buea Regional Hospital) to the Faculty of Health Sciences, Medical Research, and Bacteriology Laboratory (FHS-MRBL). The stool samples were then aliquoted into 3 containers. A container of the shared stool sample was then coded and stored in a -80 °C freezer at the Infectious Disease Laboratory, Faculty of Health Sciences, University of Buea for DNA extraction and 16sRNA sequencing

### Blood sample collection

Venous blood (10 mL) was also collected by venepuncture from participants (patients and controls) into 2 separate 5 mL ethylene-diamine-tetra-acetate (EDTA) vacutainer tubes. One of the tubes was used for HIV Screening and CD4^+^ T cell count, while the other tube was used for Viral load testing.

### HIV Screening

Briefly, HIV screening was done using the method described by Respess *et al*, [13]. The HIV determine test strip (Abbott Laboratories, Abbott Park, IL, USA) was labeled with the participant identification number. The protective foil cover of the strip was pulled off. Then 50 µL of plasma was collected with a pipette and applied to the absorbent pad on the test strip. One drop of chase buffer was added to the specimen pad and the specimen was allowed on the bench to run through the test strip. Reading and recording of the results were done after 15 minutes. Positive samples demonstrated two red lines, while negative samples had just one red line on the test strip

### CD4^+^ T cell count

The CD4^+^ T cell count of the study participant was determined using the method of Fonsah *et al*., [14]. The test was run according to a standardized Flow cytometric flow machine (BD Biosciences FACSCount, New Jersey, USA) following the manufacturer’s procedure. Briefly, the EDTA tube with the whole blood sample was mixed and 50 µL of whole blood was pipetted into the reagent tube labeled with the corresponding participant’s number. The tube was then capped, vortexed, and incubated for 30 minutes at room temperature (20°C–25°C) in the workstation. This was followed by uncaping each sample tube and pipetting 50 µL of a fixative solution into each tube. The tubes were recapped and vortexed upright before uncapping to run the sample with the Flow cytometric flow machine. Reading and recording of the results were done after the machine software message was indicated. The interpretation was done as follow: normal range 500 – 1,500 cells/µL, while below 500 cells/µL was considered as low CD4 ^+^ T cell count.

### Viral Load Measurement

HIV viral load was done using the method described by Neogi *et al*.,[15]. The viral load of each sample was measured using the Abbott RealTi*m*e HIV-1 Qualitative (Abbott Molecular Inc, Des Plaines, IL, USA) assay following the manufacturer’s instructions. Dried blood spot strips were prepared from freshly drawn whole blood after storage at room temperature for up to 6 hours, by spotting 50uL of the whole blood onto a Whatman 903 filter paper (3 spots per card). Briefly, filter papers were air-dried overnight at room temperature and stored at 4°C in a plastic sealed bag with a silica desiccant until they were processed. Dried blood spot viral load was measured as follows: two blood spots from the same patient were punched out using a sterile puncher, and placed into 1.7 ml of Lysis buffer provided with the Abbott sample preparation system (m2000sp) in 50 ml sealed conical tubes. The tubes were incubated at room temperature for 2 h, with intermittent mixing. RNA was extracted manually from the lysate according to the standard HIV-1 RNA 1.0 ml extraction protocol using the Abbott RNA sample preparation system. The viral load was measured from the extracted RNA using the “m2000 DBS HIV-1 RNA ‘open-mode’ protocol” (Abbott Molecular Inc, Des Plaines, IL, USA). Reading and recording of the viral load values were stratified into three levels: *(i)* VL 2.17 to 3 log_10_ copies/ml (corresponding to 1000 copies/ml), *(ii)* VL >3 to 3.7 log_10_copies/ml, (1000-about 5000 copies/ml), and *(iii)* VL >3.7 log_10_ copies/ml (corresponding to approximately 5000 copies/ml).

### Extraction and Purification of Microbiota DNA from the Stool Sample

Extraction of bacterial DNA procedure was done using the method described by Shantelle *et al*. [16] with the ZymoResearch DNA MiniPrep Extraction kit (Zymo, Irvine, CA, USA). The Stored (−80 ^0^c) fecal sample (200mg) was purposively selected (**Table 1**) and added to a ZR Bashing bead lysis tube. Then 750 μL of ZymoBIOMICS lysis solution was added to the tube and cap tightly. The tube was then secured in a bead beater fitted with a 2 mL tube holder assembly and it was processed at maximum speed for 20 minutes. The beaten ZR Bashing Bead lysis tube was then centrifuged at ≥ 10,000 × g for 1 minute. The supernatant (400 μL) was transferred to the Zymo-Spin III-F filter in a collection tube and centrifuged at 8,000 × g for 1 minute. Then the used Zymo-Spin III-F filter was discarded. Binding preparation was done by adding 1,200 μL of ZymoBIOMICS DNA binding buffer to the filtrate in the collection tube and the tube was mixed thoroughly. The mixture (800uL) from the binding step was transferred to a Zymo-spin IIC-Z column in a collection tube and centrifuged at 10,000 × g for 1 minute. The flow-through from the collection tube was discarded and the aforementioned step was repeated.

**Table 1:** Characteristics of Fecal samples purposively selected for 16S rRNA sequencing.

| Codes | Type of samples | No. Extracted |
| --- | --- | --- |
| N.01 | HIV negative extracted stool samples | 5 |
| N.02 | HIV negative extracted stool samples | 5 |
| N.03 | HIV negative extracted stool samples | 5 |
| PT.04 | HIV-TB co-infected extracted stool samples | 4 |
| PT.06 | HIV-TB co-infected extracted stool samples | 3 |
| P.05 | HIV positive extracted stool samples | 4 |
| P.07 | HIV positive extracted stool samples | 4 |
| P.08 | HIV positive extracted stool samples | 4 |
| PN.09 | HIV positive treatment-naïve extracted stool samples | 3 |
| PN.010 | HIV positive treatment-naïve extracted stool samples | 3 |

The Zymo-spin IIC-Z column containing the settled contents was then inserted into a new collection tube. ZymoBIOMICS DNA wash buffer 1 (400 μL) was then added to the Zymo-spin IIC-Z column in the new collection tube and centrifuged at 10,000 × g for 1 minute. Then the flow-through was discarded. ZymoBIOMICS DNA wash buffer 2 (700 μL) was then added to the Zymo-spin IIC-Z column in the new collection tube and centrifuged at 10,000 × g for 1 minute. The flow-through was then discarded. ZymoBIOMICS DNA wash buffer 2 (200 μL) was then added to the Zymo-spin IIC-Z column containing the settled contents in the new collection tube and centrifuged at 10,000 × g for 1 minute. The Zymo-spin IIC-Z column was transferred to a clean 1.5mL micro-centrifuge tube and 100 μL of ZymoBIOMICS DNase/RNase free water was added directly to the column matrix and incubation was done for 1 minute. Then centrifugation was carried out at 10,000 × g for 1 minute to elute the DNA. Zymo-spin III-HRC filter was placed in a new collection tube and 600 μL of ZymoBIOMICS HRC prep solution was added and centrifuged at 8,000 × g for 3 minutes.

The eluted DNA was then transferred to the prepared Zymo-spin III-HRC filter in a clean 1.5 mL micro-centrifuge tube and centrifuged at 16,000 × g for 3 minutes. The extracted DNA was purified with Clean and Concentrator-25 columns (Zymo, Irvine, CA, USA) as per the manufacturer’s directives. Isolated DNA was stored at -80 °C until analyzed.

The DNA samples were prepared for targeted sequencing with the *Quick*-16S™ NGS Library Prep Kit (Zymo Research, Irvine, CA). The primers were custom-designed by Zymo Research to provide the best coverage of the 16S gene while maintaining high sensitivity. The primer sets used was *Quick*-16S™ Primer Set V3-V4 (Zymo Research, Irvine, CA). The sequencing library was prepared using an innovative library preparation process in which PCR reactions were performed in real-time PCR machines to control cycles and therefore limit PCR chimera formation. The final PCR products were quantified with qPCR fluorescence readings and pooled together based on equal molarity. The final pooled library was cleaned up with the Select-a-Size DNA Clean & Concentrator™ (Zymo Research, Irvine, CA), then quantified with TapeStation® (Agilent Technologies, Santa Clara, CA) and Qubit® (Thermo Fisher Scientific, Waltham, WA).

### Control Samples

The ZymoBIOMICS® Microbial Community DNA Standard (Zymo Research, Irvine, CA) was used as a positive control for each targeted library preparation. Negative controls (i.e. blank extraction control, blank library preparation control) were included in each run to assess the level of bioburden carried by the wet-lab process.

#### Sequencing

The final library was sequenced on Illumina® MiSeq™ with a v3 reagent kit (600 cycles) by the Zymo Research Corporation in USA. The sequencing was performed with >10% PhiX spike-in.

### Bioinformatics Analysis

Unique amplicon sequences were inferred from raw reads using the Dada2 pipeline [17]. Chimeric sequences were also removed with the Dada2 pipeline. Taxonomy assignment was performed using Uclust from Qiime v.1.9.1. Taxonomy was assigned with the Zymo Research Database, a 16S database that is internally designed and curated, as reference.

Composition visualization, alpha-diversity, and beta-diversity analyses were performed with Qiime v.1.9.1 [18]. The taxonomy that has significant abundance among different groups was identified by LEfSe [19] using default settings. Other analyses such as heatmaps, Taxa2SV Deomposer, and PCoA plots were performed with internal scripts.

Diversity was analyzed following a previous method described by Landro *et al*. [20] in which the Shannon index (*H*′) measured the average degree of uncertainty in predicting what species an individual is chosen at random from a collection of *S* species and *N* individuals will belong. The value increases as the number of species increases and as the distribution of individuals among the species become even. Meanwhile, Simpson’s index (*D*) Indicates species dominance and reflects the probability of two individuals that belong to the same species being randomly chosen. It varies from 0 to 1 and the index increases as the diversity decreases. And lastly, the Chao1 richness estimator is a Non-parametric estimator that calculates the minimal number of OTUs present in the sample

### Data availability

The raw sequencing data was zipped and can be accessed at: https://epiquest.s3.amazonaws.com/epiquestzr2768/CMCDFQUJAMKPZKAFWSY3FJLBJDRTBYVE/rawdata/zr2768.rawdata.190904.zip

## RESULTS

### Sociodemographic and clinical Characteristics of Study Partticipants

The characteristics of the study participants purposively selected from the cohort are shown in **Table 2**. A total of 15 HIV-negative individuals and 25 HIV-infected persons were enrolled in the study. The HIV-infected cases were further stratified based on their treatment status. Most of the study participants were females 28 (70%). The age ranges were equally represented. With regards to food and drinks consumed, all participants that were purposively selected were on Energy + body-building+ protective foods (100%), and non-alcoholic drinks (100%).

**Table 2:**
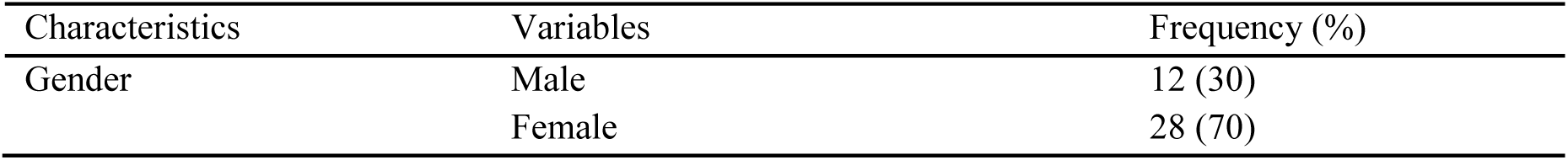
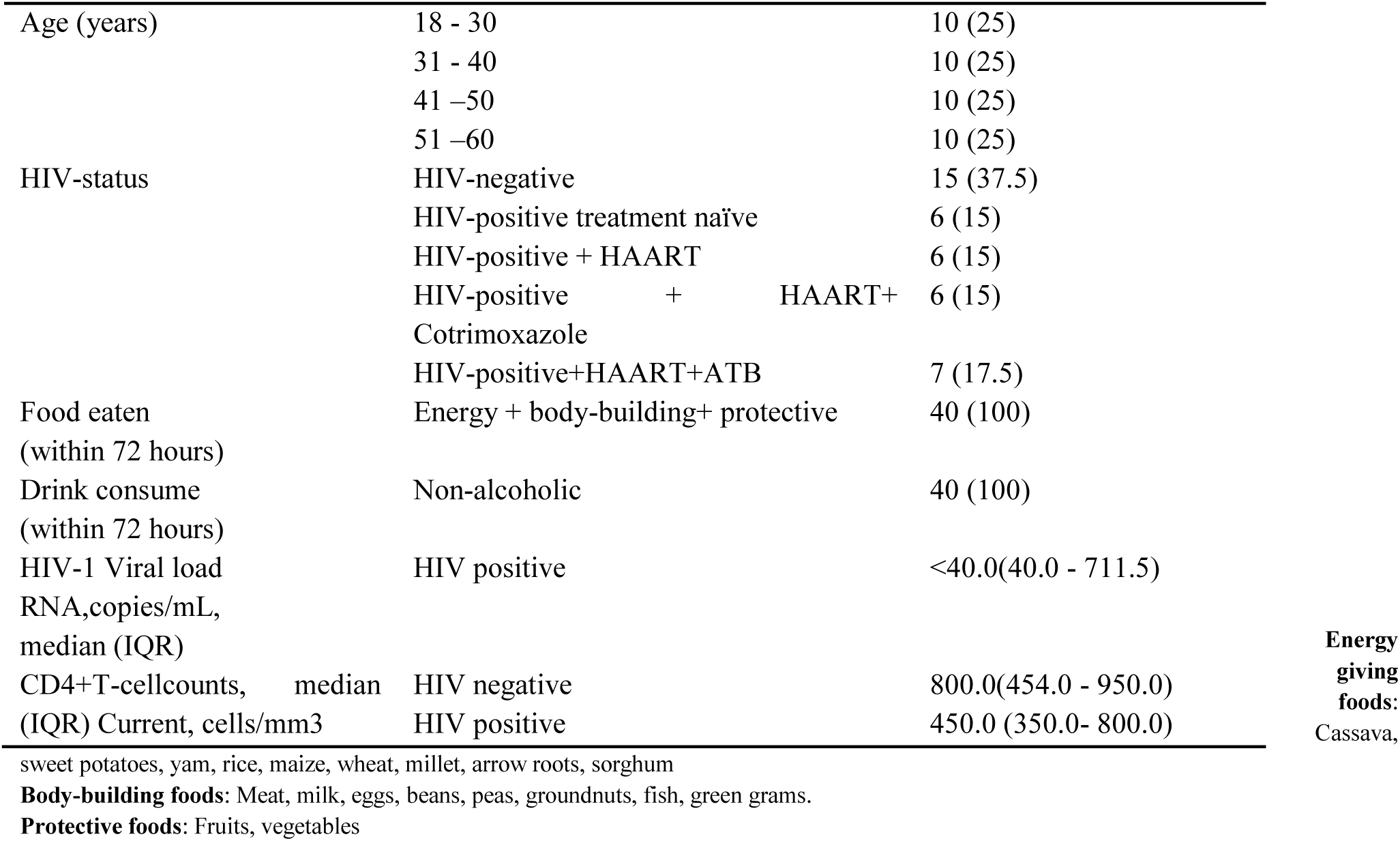
Sociodemographic and clinical Characteristics of the study participants in Cameroon.

The mean HIV viral load was <40.0(40.0 - 711.5) RNA, copies/mL, while the mean CD4+T-cellcounts was 800.0(454.0 - 950.0) cells/mm3 and 450.0 (350.0-800.0) cells/mm3 for HIV-negative and HIV-infected respectively.

### Gut Bacterial Microbiome Composition in the study population

Summarily we identified, characterized, and compared microbiome communities between HIV-positive and HIV-negative individuals using culture-independent techniques. The complete data are shown in Appendixes one and two. Taxonomy of the microbiome communities ranged from two kingdom (Archea and Bacteria), and eight Phylum [Firmicutes (44.7%), Bacteroidetes (43.7%), Proteobacteria (8.7%), Actinobacteria (1%), Fusobacteria (0.2%), Euryarchaeota (0.01%) Synergistetes (0.01%), Verrucomicrobia (0.01%) and unclassified phylum (1,7%)]. Totally 347 species were characterized. Notably at the level of Family, Genus, and Species, there were unknown/unclassified microbial communities 2/39, 11/102, and 7/347 respectively. Comparing the total taxonomic community structure between the two groups (HIV-positive and HIV-negative) showed variation at all levels from Kingdom to Specie (**Table 3** and Appendix one).

**Table 3:**
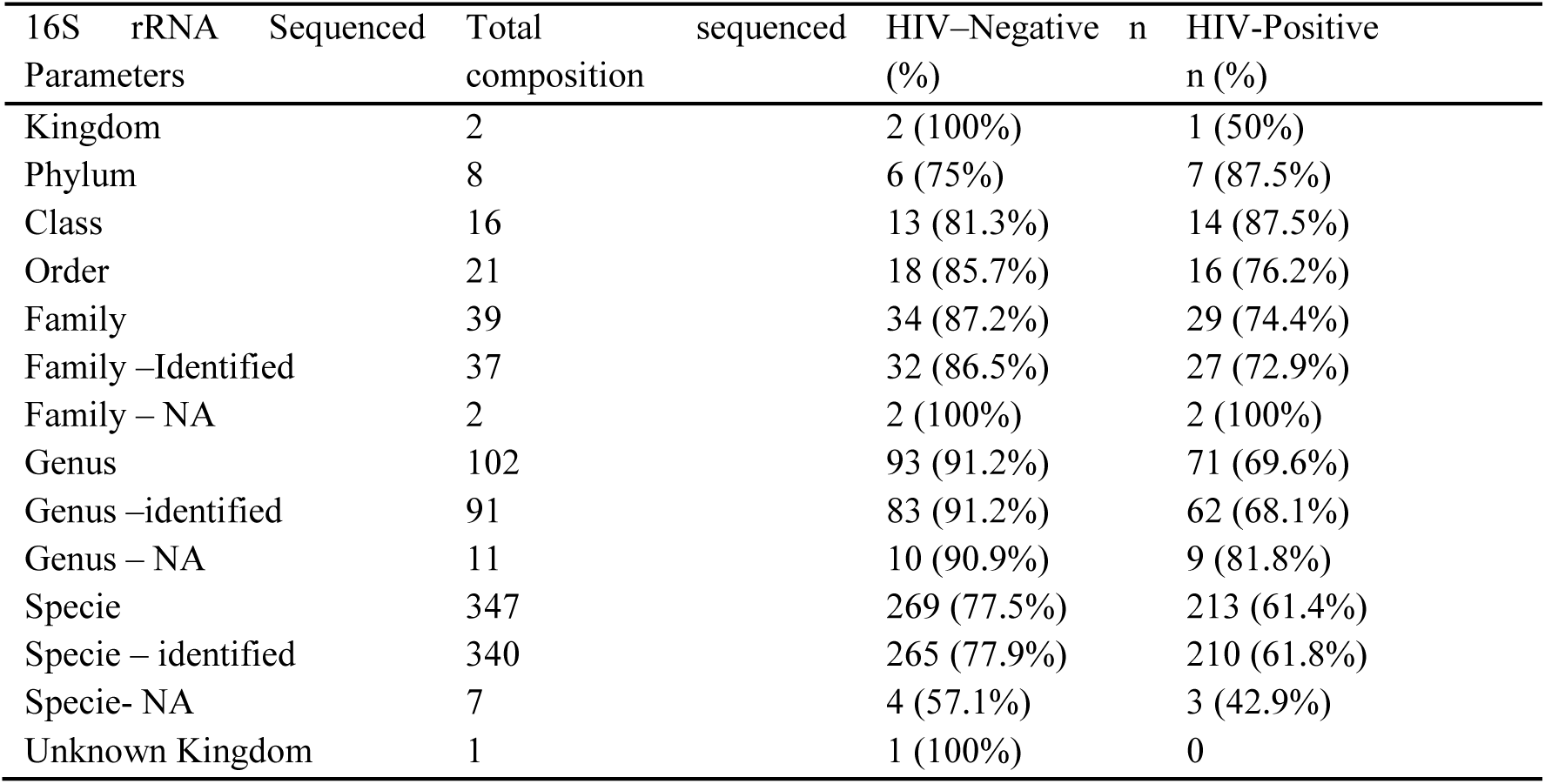
Summarized phylogenetic classification of gut microbiome in study participants.

**Table 3:** Comparing HIV-infected and HIV-negative gut microbiome strains identified.

| Phylum | Gram stain | Strain (n) | HIV-negative (%) | HIV-infected (%) |
| --- | --- | --- | --- | --- |
| Actinobacteria | + | 16 | 13 (81.3) | 5 (31.3) |
| Bacteroidetes | + | 67 | 29 (43.3) | 50 (74.6) |
| Firmicutes | + | 220 | 175 (79.6) | 141 (64.1) |
| Fusobacteria | - | 4 | 4 (100) | 1 (25) |
| Proteobacteria | - | 36 | 31 (86.1) | 12 (33.3) |
| Synergistetes | - | 1 | 0 (0) | 1 (100) |
| Verrucomicrobia | - | 1 | 0 (0) | 1 (100) |
| Euryarchaeota | + | 1 | 1 (100) | 0 (0) |

### Taxonomy abundance variation of OTU for the 50 most abundant microbial strains

In HIV-negative individuals, the most abundant fecal microbiota at the genus and species levels included: *Collinsella aerofaciens, Bacteroides uniformis, Bacteroides vulgatus, Parabacteroides merdae, Prevotella copri, Streptococcus salivarius-vestibularis, Clostridium celatum, Anaerostipes hadrus, Blautia sp32009, Blautia sp32056, Blautia wexlerae, Coprococcus comes-sp32193, Dorea formicigenerans, Dorea longicatena, Eubacterium hallii, Eubacterium rectal, Fusicatenibacter saccharivorans, Roseburia faecis-sp33781, Roseburia inulinivovans, Romboustsia ilealis, Faecalibacterium prausnitzii, Ruminiclostridium sp34909, Ruminococcus bromii, Subdoligranulum sp35582, Veillonella parvula, Lachnoclostridium sp32380-sp32437, Lachnoclostridium sp32400 and Escherichia-shigella coli* (**Figure 1**).

**Figure 1:**
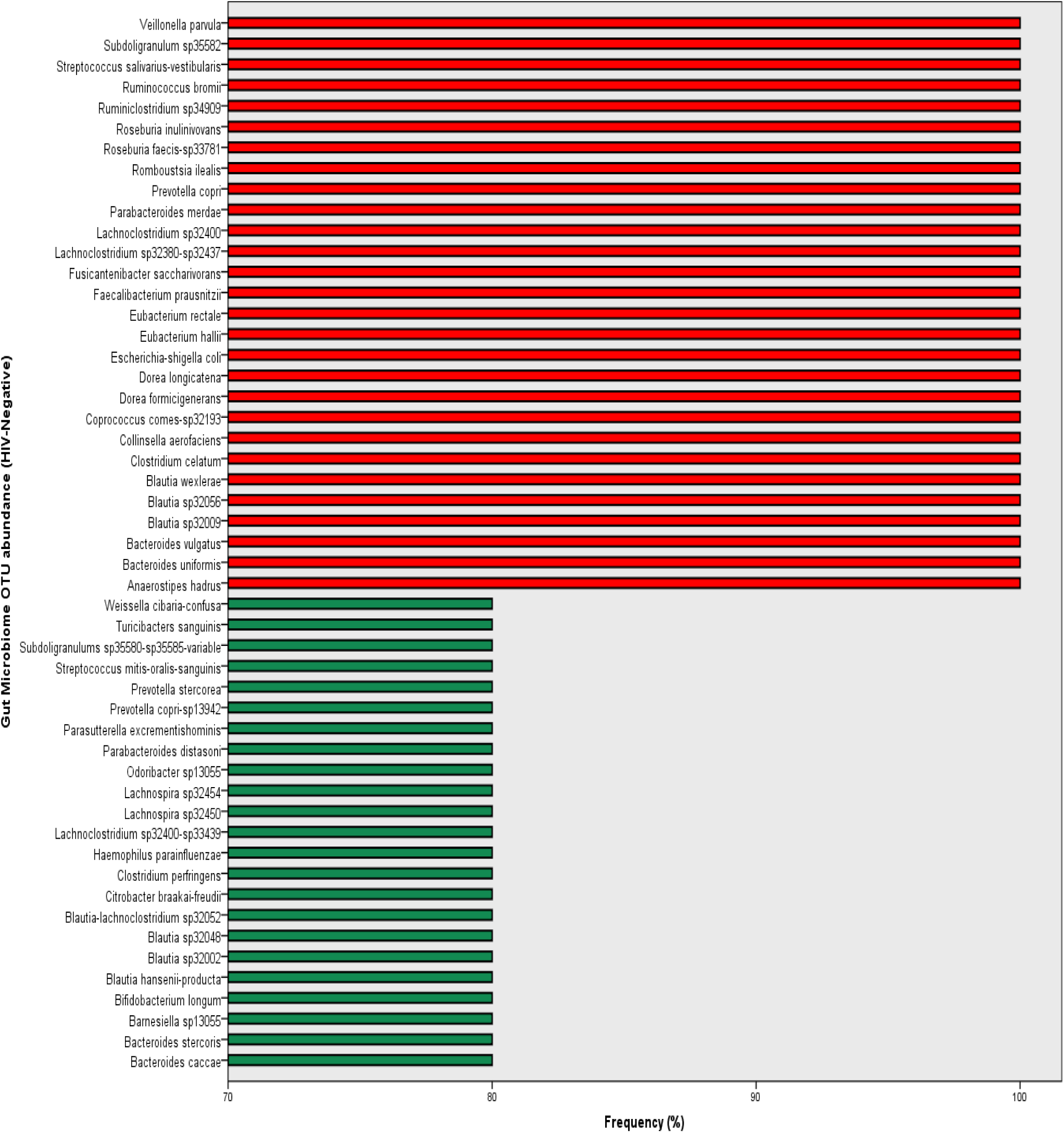
Bar chart depicting most abundant gut microbiome strains amongst HIV-negative individuals in Cameroo**n**

Also findings from the study revealed abundant unidentified strains among the HIV-negative individuals originating mostly from the Family Lachnospiraceae and Ruminococcaceae (**Figure 2**). In patients with HIV, the most abundant faecal microbiota at the genus and species levels include: *Parabacteroides distasonis, Prevotella copri, Blautia wexlerae, Lachnoclostridium sp32400, Faecalibacterium prausnitzzi, Bacteroides vulgatus, Prevotella copri-sp13942, Megamonas funiformis, Anaerostipes hadrus, Coprococcus comes-sp32193, Lachnoclostridium sp32343-sp32393-sp32423, Faecalibacterium sp34558, Subdoligranulum sp35380-sp35585, Prevotella stercorea, Blautia sp32056, Eubacterium hailli, Ruminiclostridium sp34921-sp34937, Bilophila wadsworthia, Escherichia-shigella coli, Senegalimassilia anaerobia, Bacteroides fragilis-ovatus, Bacteroides uniformis, Sutterella wadsworthensis, Holdemanella biformis, Holdemanella sp36738, Subdoligranulum sp35580-sp35585-variabile, Romboutsia ilealis, Roseburia intestinalis, Blautia sp32002, Blautia-lachnoclostridium sp32052-sp32410, Fusicatenibacters saccharivorans and Lachnospira sp32454* (**Figure 3**). Also, the majority of the unidentified gut microbiome strains among HIV-positive were noted mostly from Prevotellaceae and Lachnospiraceae families (**Figure 4**).

**Figure 2:**
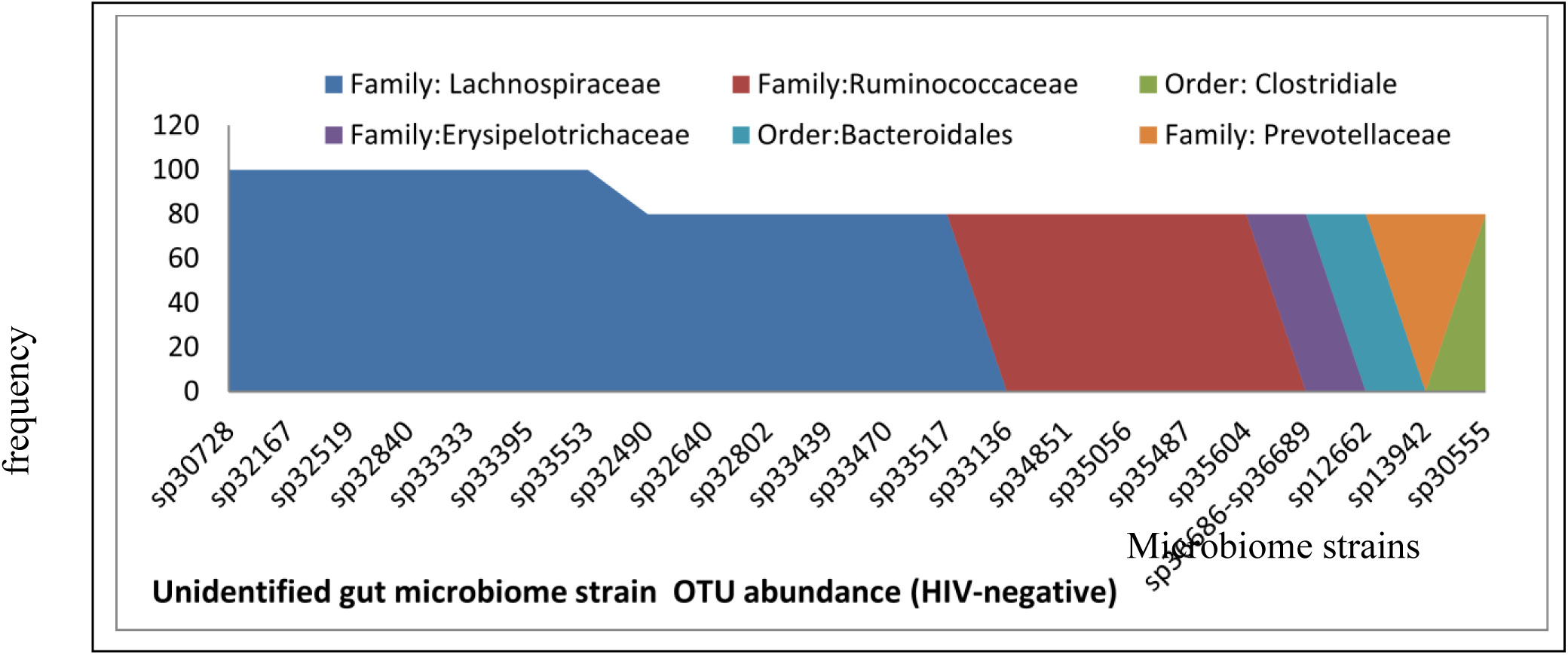
Bar plots depicting unclassified gut microbiome strains amongst HIV-negative individuals in Cameroon

**Figure 3:**
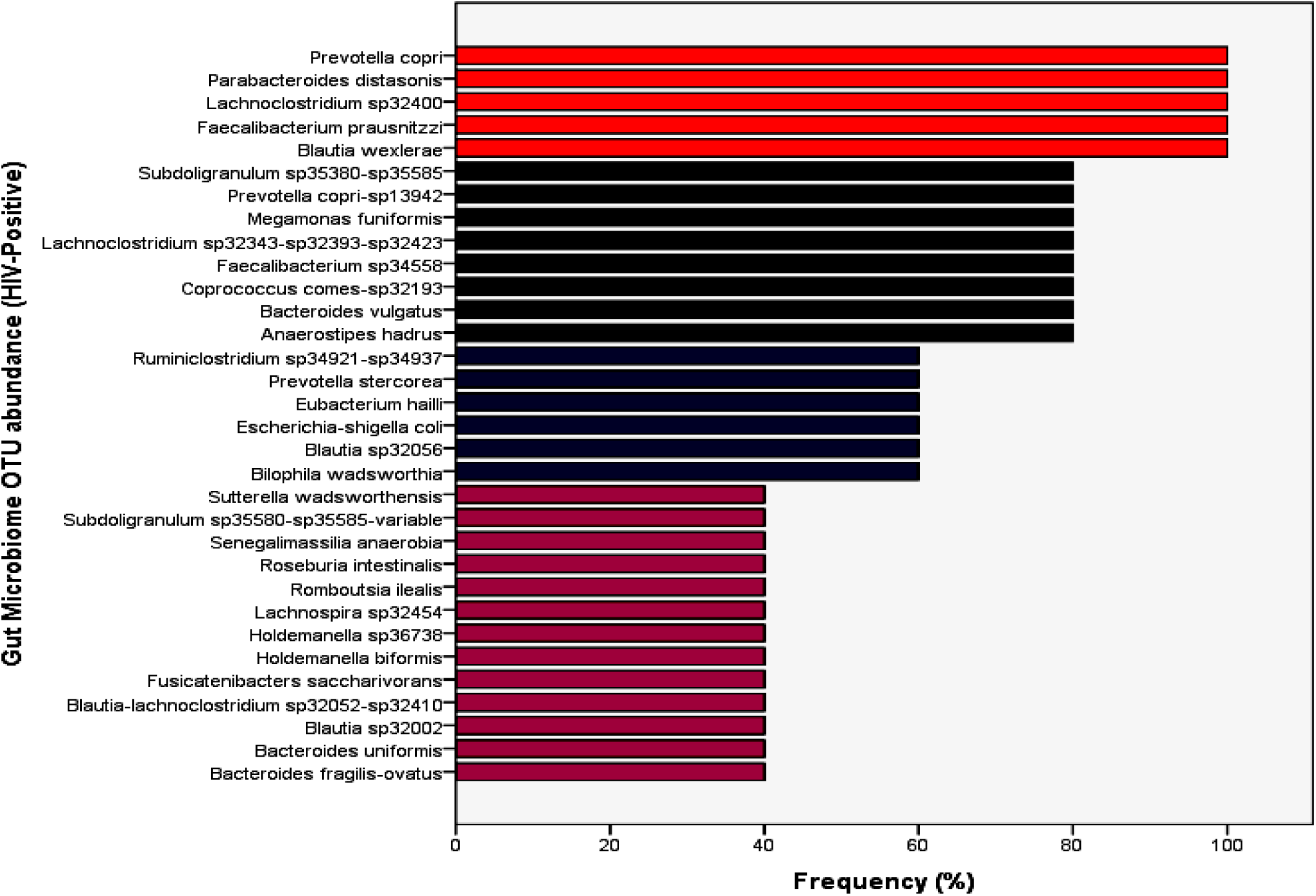
Bar chart depicting gut microbiome strains amongst HIV-positive individuals in Cameroon

**Figure 4:**
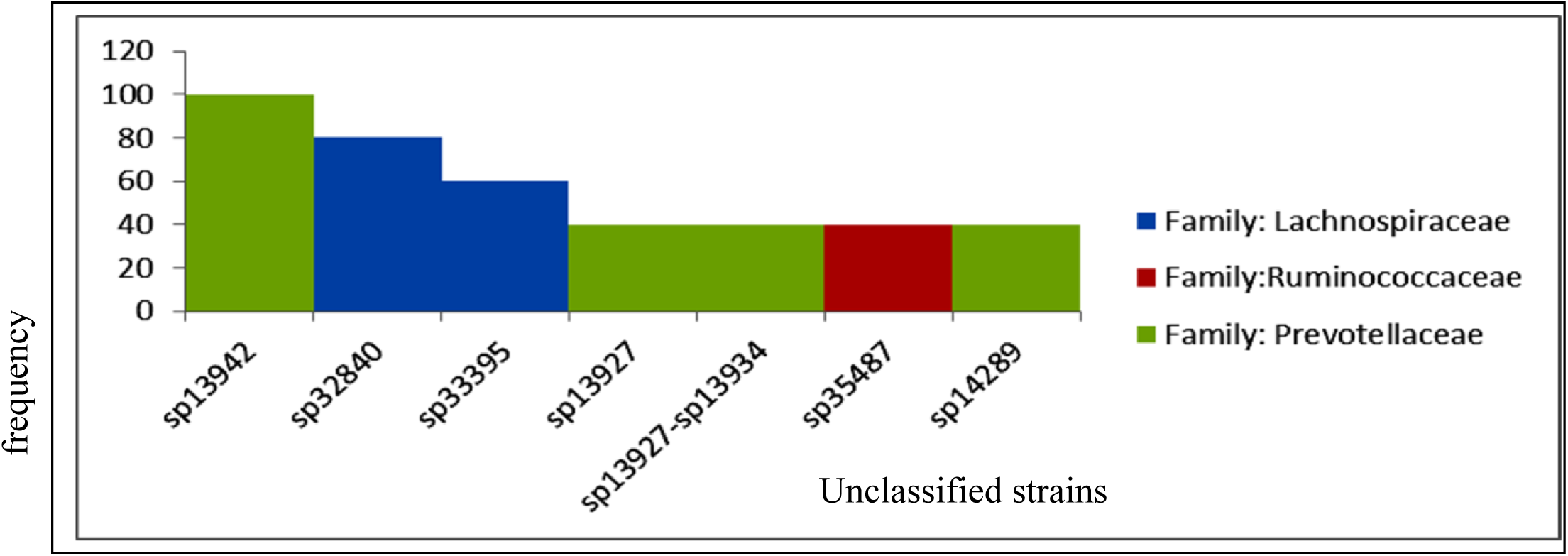
Bar plots depicting unclassified gut microbiome strains amongst HIV-positive individuals Cameroon

#### 4.2.3 Dysbiotic pattern amongst HIV-negative Individuals Compared to HIV-positive individuals with or without treatment on ARV/or Cotrimoxazole prophylaxis

The Operational Taxonomy Unit (OTU) of the microbial community when compared between HIV-positive and HIV-negative individuals showed dysbiosis between the different taxa (**Table 3**). HIV patients had significantly reduced microbial richness (p < 0.05) compared to HIV-negative individuals (**Figure 5**). However, some microbial communities including *Prevotella copri, Blautia wexlerae, Faecalibacterium prausnitzii, Ruminiclostridium sp34909, Lachnoclostridium sp32400, Parabacteroides distasoni, Prevotella copri-sp13942, and Blautia sp32048* shared the same OTU abundance between the two groups.

**Figure 5:**
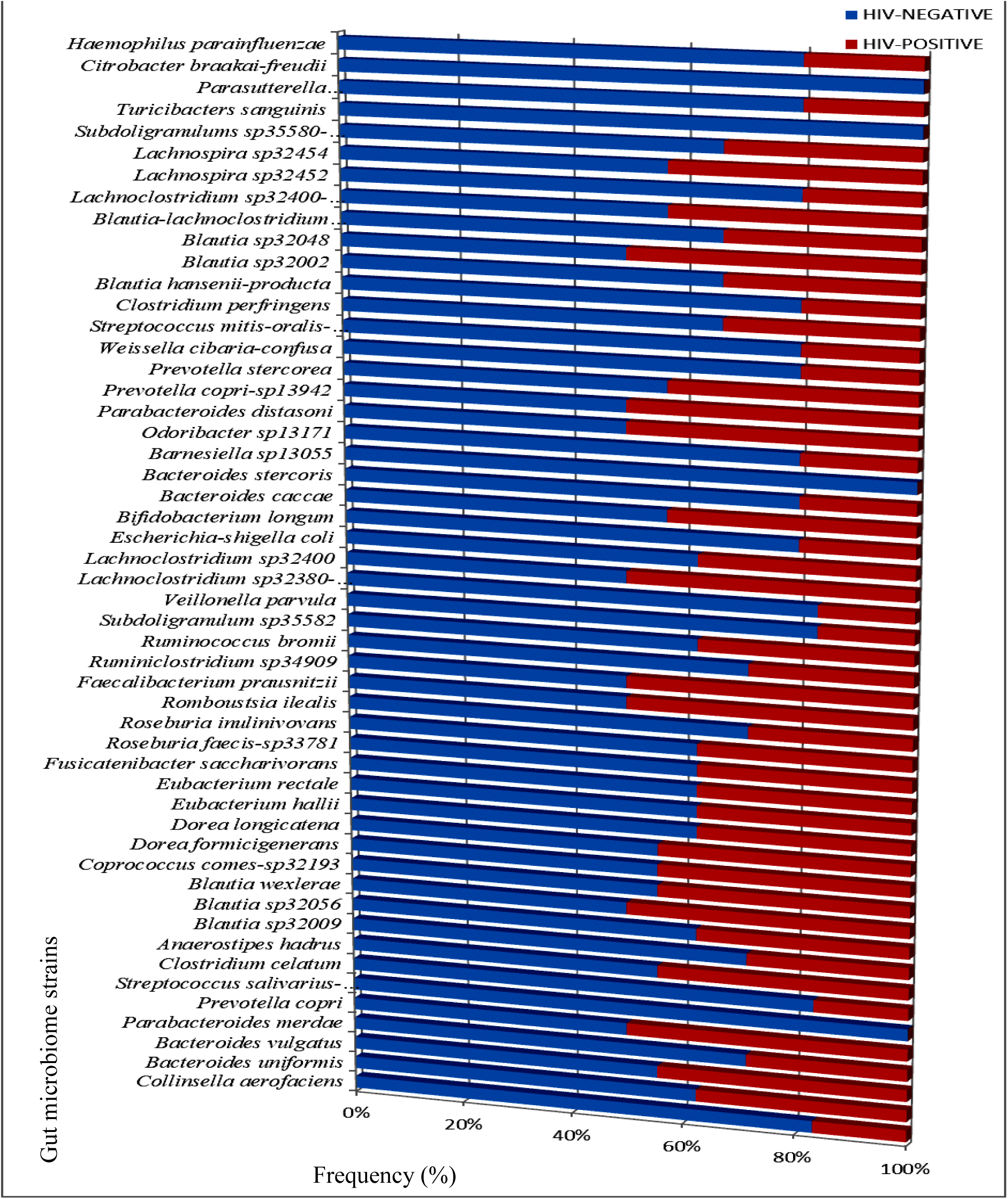
Gut microbiome dysbiosis profiles at the genus level between HIV-positive and HIV-negative individuals

### Unique Gut Microbiome Strains/Oligotype Circulating Amongst the Study participants

Findings from this study showed that 55 species of the 347 gut microbiome strains identified contained more than one unique sequence. The sequences were distributed among HIV-positive and HIV-negative individuals in an uneven manner. Some of the unique sequences with increased abundance among HIV-negative individuals included *Bifidobacterium (seq67 Bifidobacterium adolescentis-faecale* and *seq159 Bifidobacterium longum)* (**Figure 6**), *Fusicatenibacter (seq75 Fusicatenibacter saccharivorans)* (**Figure 7**), *Escherichia-Shigella coli (seq30 Escherichia-Shigella;coli)* (**Figure 8**), *Collinsella (seq 40 Collinsella aerofaclens* and *Romboutsia (seq27 Romboutsia ilealis)*.

**Figure 6:**
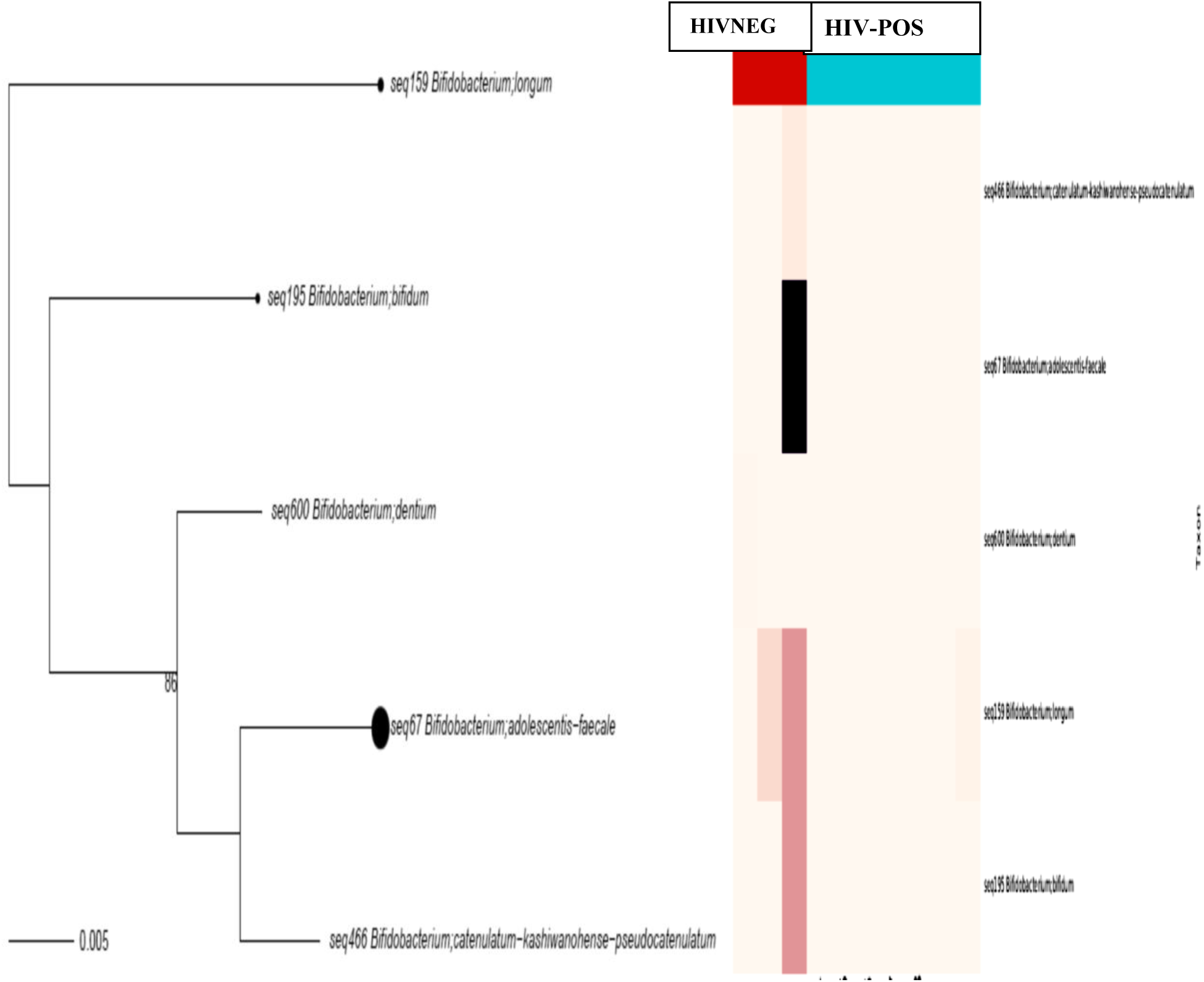
*Bifidobacterium* Unique sequence. Heat map illustrating seq159 *Bifidobacterium longum* and seq67 *Bifidobacterium adolescentis–faecale* unique amplicon present only among HIV-negative individuals and absent among HIV-positive individuals

**Figure 7:**
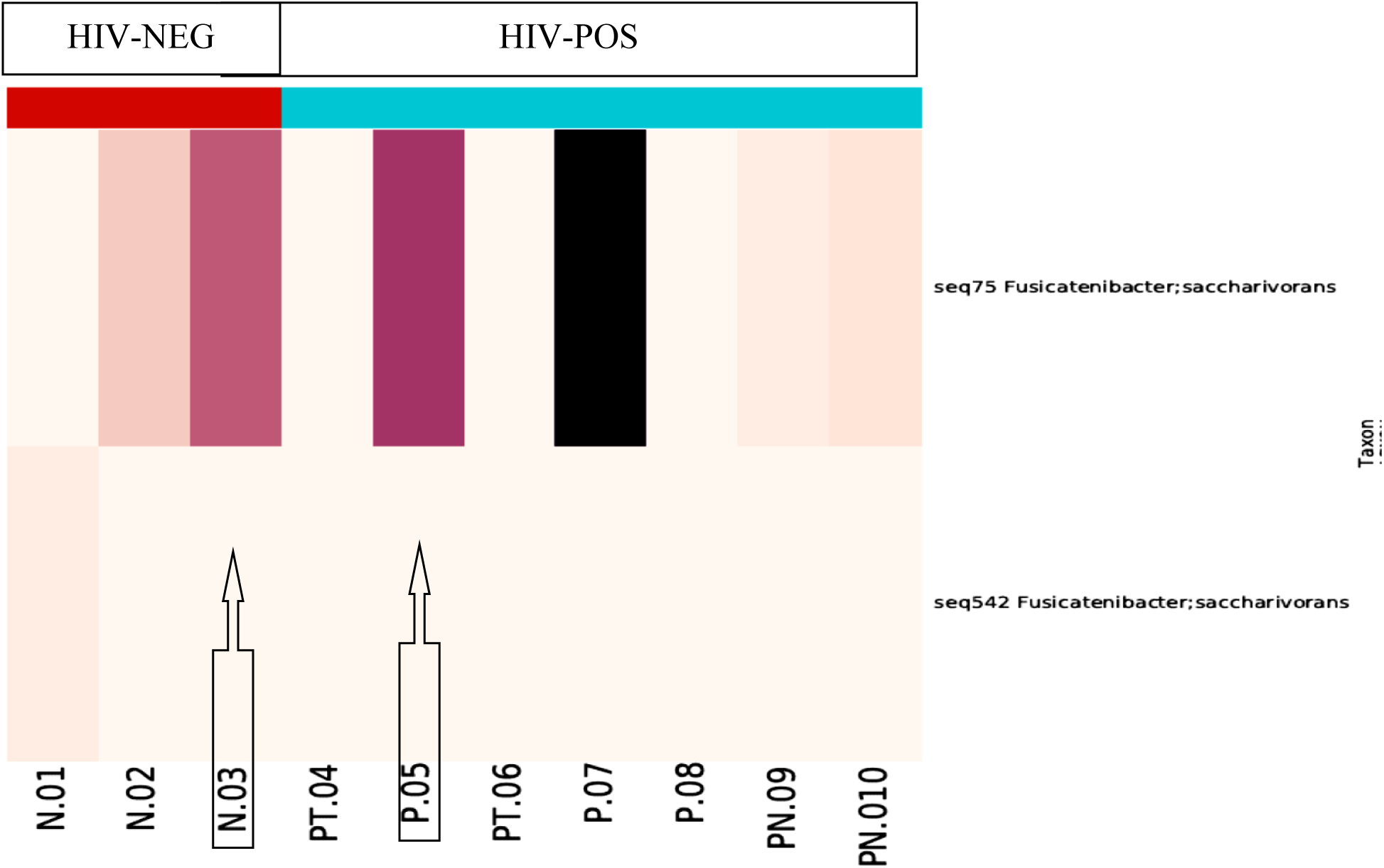
*Fusicatenibacter saccharivorans* unique sequence amplicons. Heat map illustrating *Fusicatenibacter saccharivorans* seq75 absent only in HIV/TB coinfected individuals and present in both HIV-negative and HIV-positive without TB infection.

**Figure 8:**
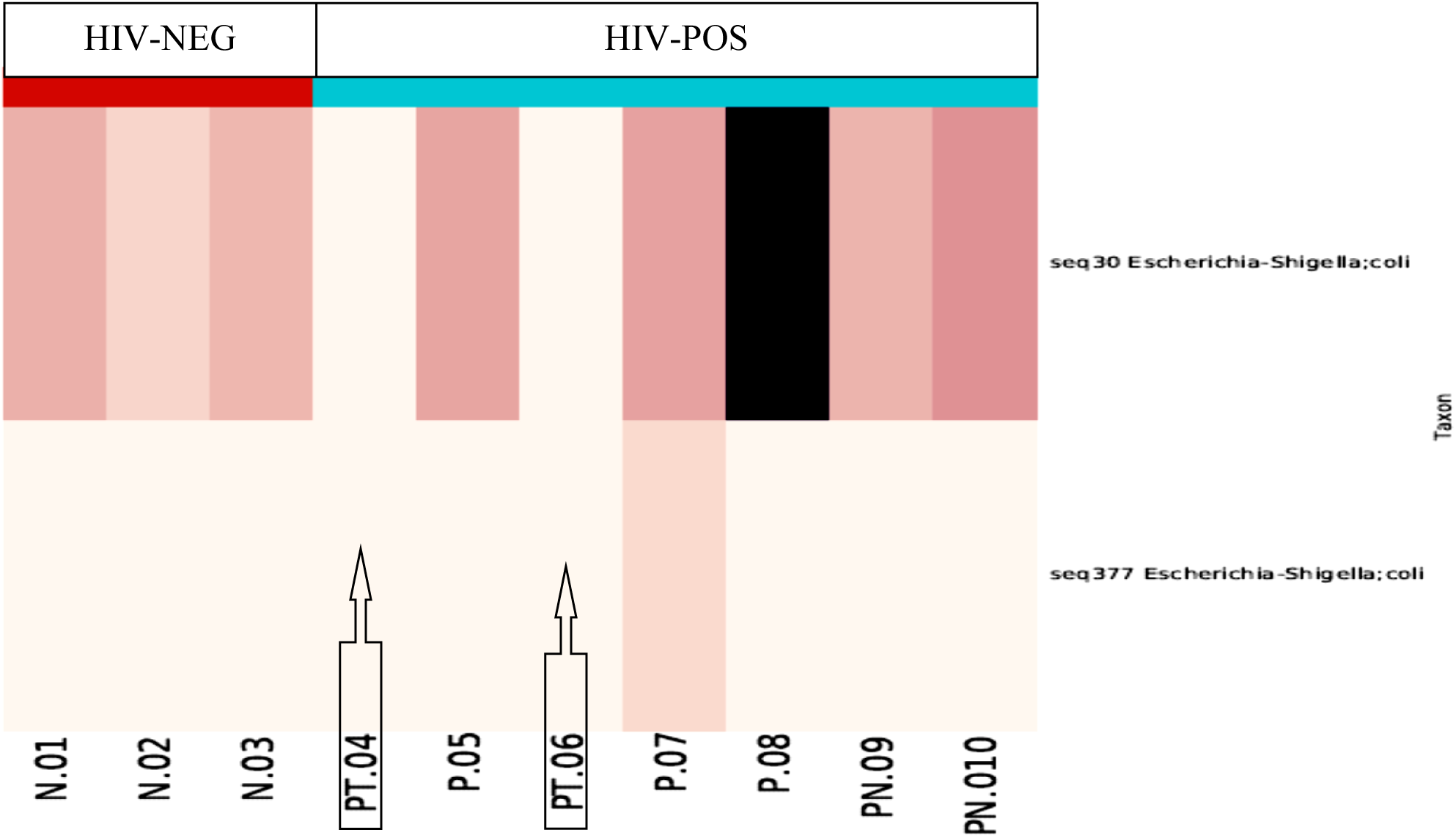
*Escherichia-Shigella coli* unique sequence amplicon. Heat map illustrating *Escherichia-Shigella coli* seq 30. absent only in HIV/TB coinfected individuals and present in both HIV-negative and HIV-positive without TB infection.

Also among HIV infected individuals, some unique Taxons amplicon sequence, were significantly increased among HIV patients on HAART treatment even though they were on cotrimoxazole medication. The increased unique amplicon sequence OTU observed included *Subdoligranulum (seq 48, seq114, seq 201 and seq 396)*(**Figure 9**), *Lachnospira (seq 76, seq 184, and seq389), Megamonas funiformis (seq 2, seq12, seq 16, seq 21 and seq 22)* (**Figure 10**), *Catenibacter mitsoukai (seq 152, seq 234, seq 254 and seq 353)* (**Figure 11**), *Bacteroides (seq 7, seq 8, seq 47, seq 74, seq 79, seq 77, seq 83, seq 87, seq 113 and unidentified species)* (**Figure 12**), *Ruminiclostridium (seq220 and seq185)*(**Figure 13**) *and Eubacterium rectale (seq 11 and seq 60)*

**Figure 9:**
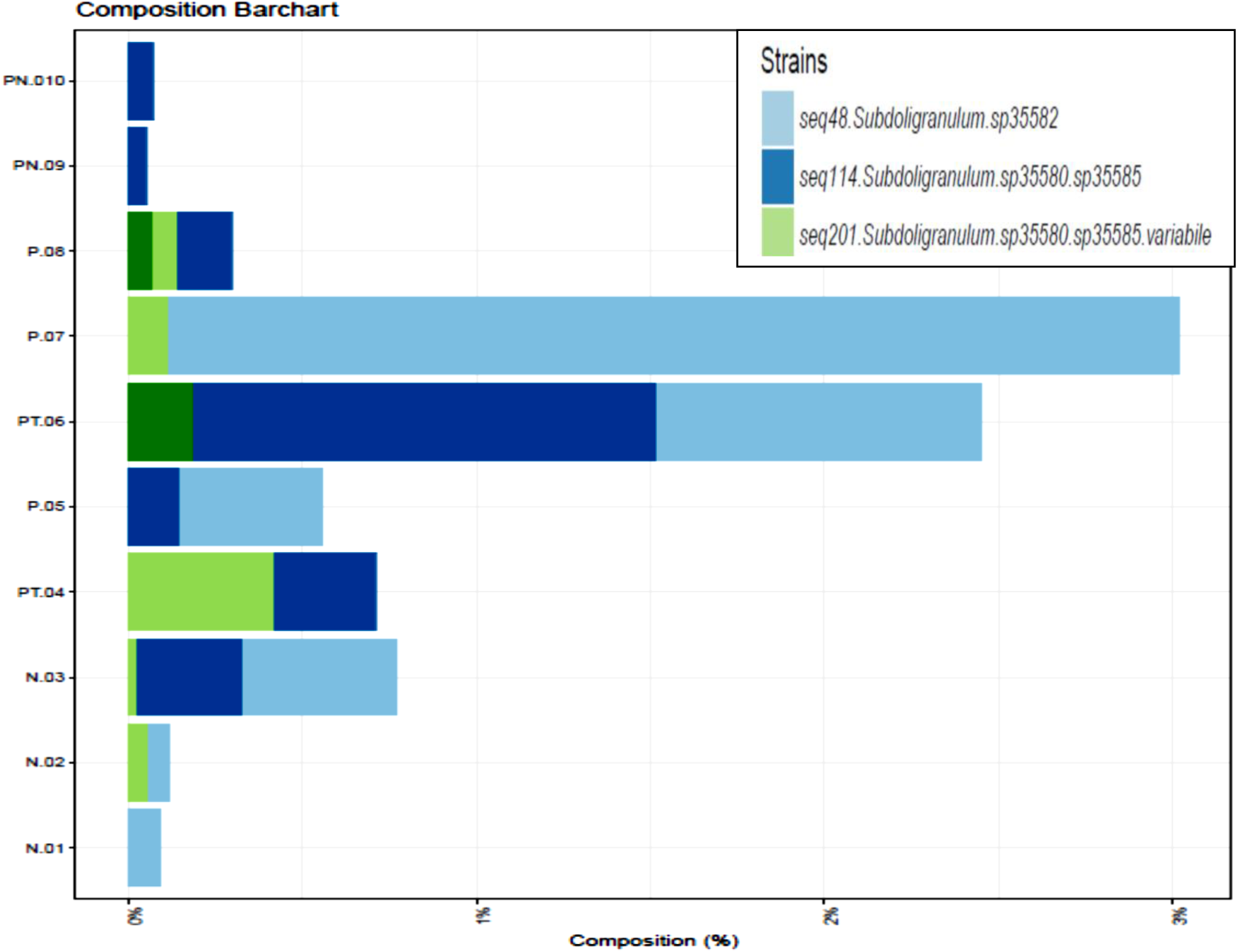
*Subdoligranulum* unique taxon amplicon. Bar charts depicting increased variation of unique sequences for *Subdoligranulum* amongst HIV-positive individuals on HAART and HIV/TB coinfected.

**Figure 10:**
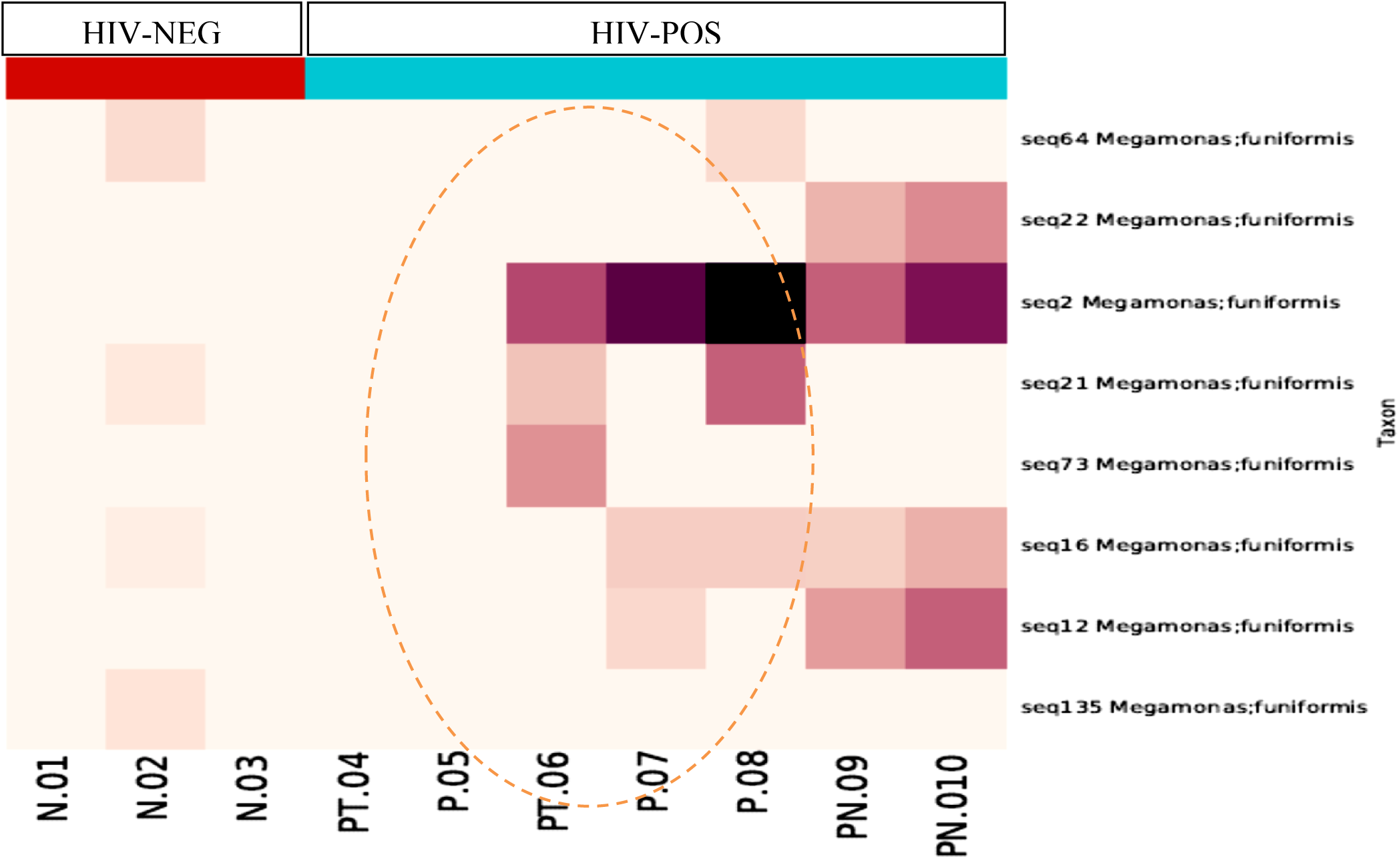
*Megamonas funiformis* unique taxon amplicon. Heat map depicting increased variation of unique sequences for *Megamonas funiformis* amongst HIV-positive and HAART naïve treatment individuals only.

**Figure 11:**
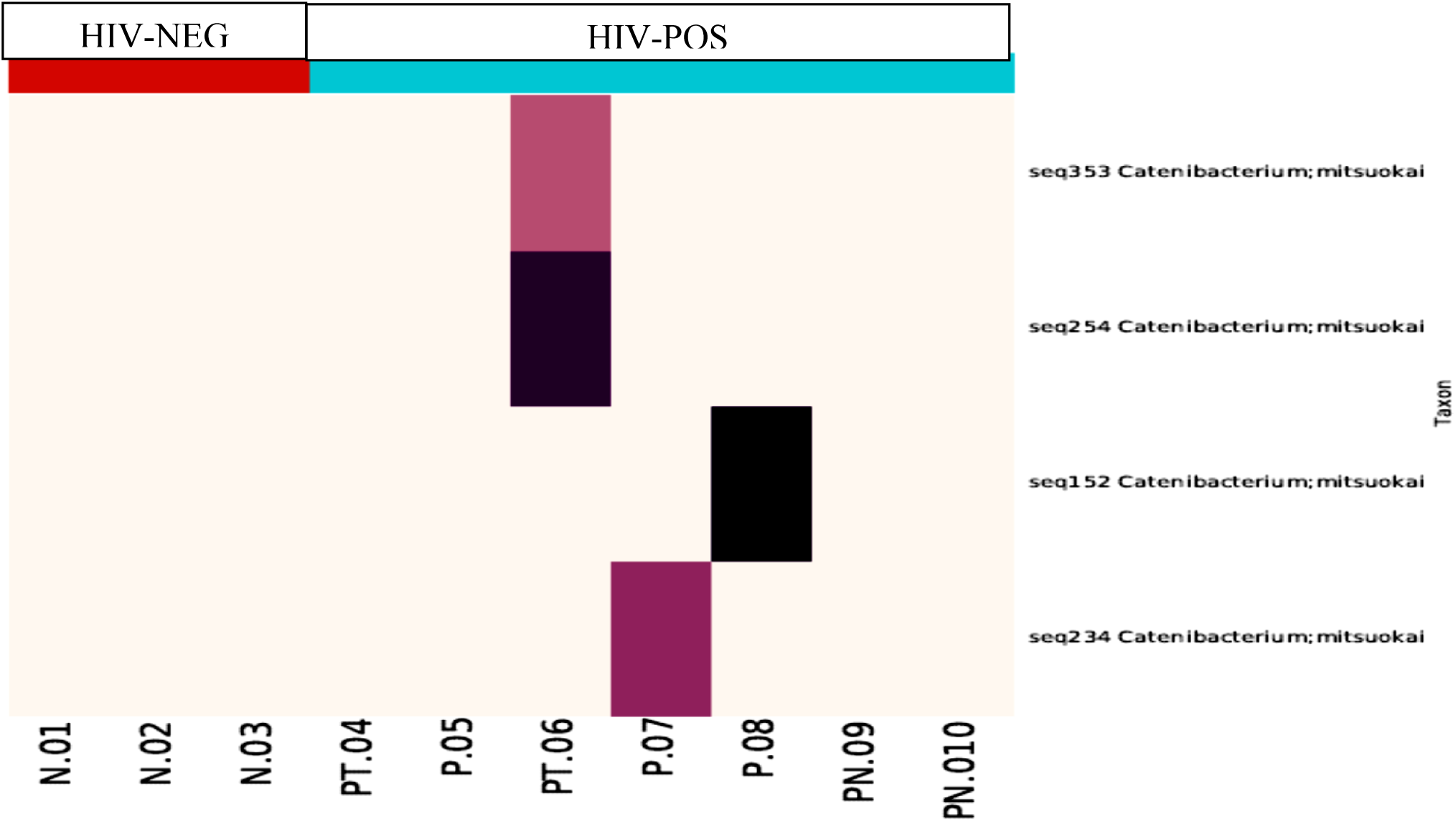
*Catenibacterium mitsuokai* unique taxon sequence. Heat map depicting increased variation of unique sequences for *Catenibacterium mitsuokai* amongst HIV-positive and HAART naïve treatment individuals only

**Figure 12:**
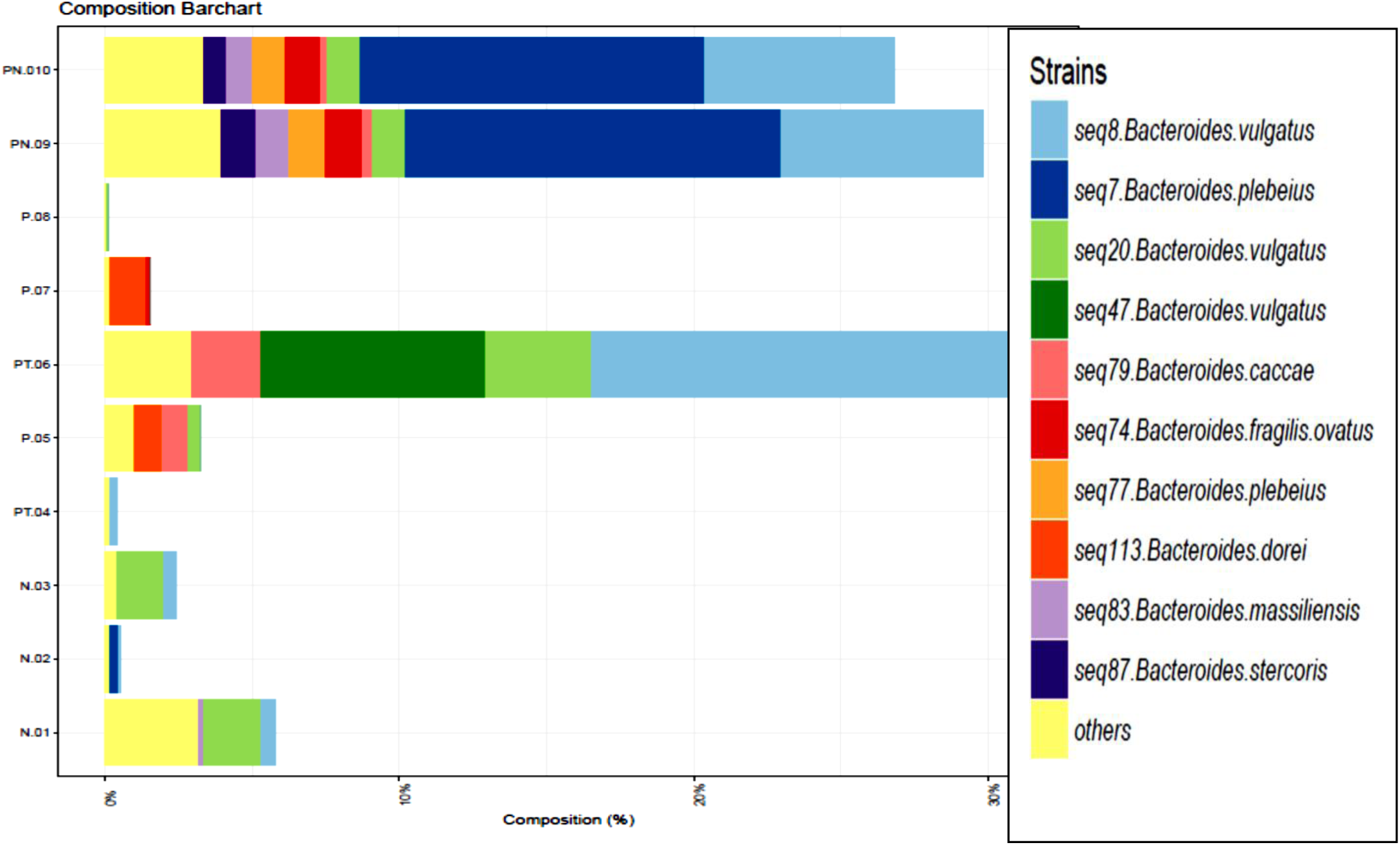
*Bacteroides* unique amplican sequence distribution. Bar charts depicting increased variation of unique sequences for *Bacteroides strains* amongst HIV-positive treatment naïve individuals and HIV/TB coinfected individuals.

**Figure 13:**
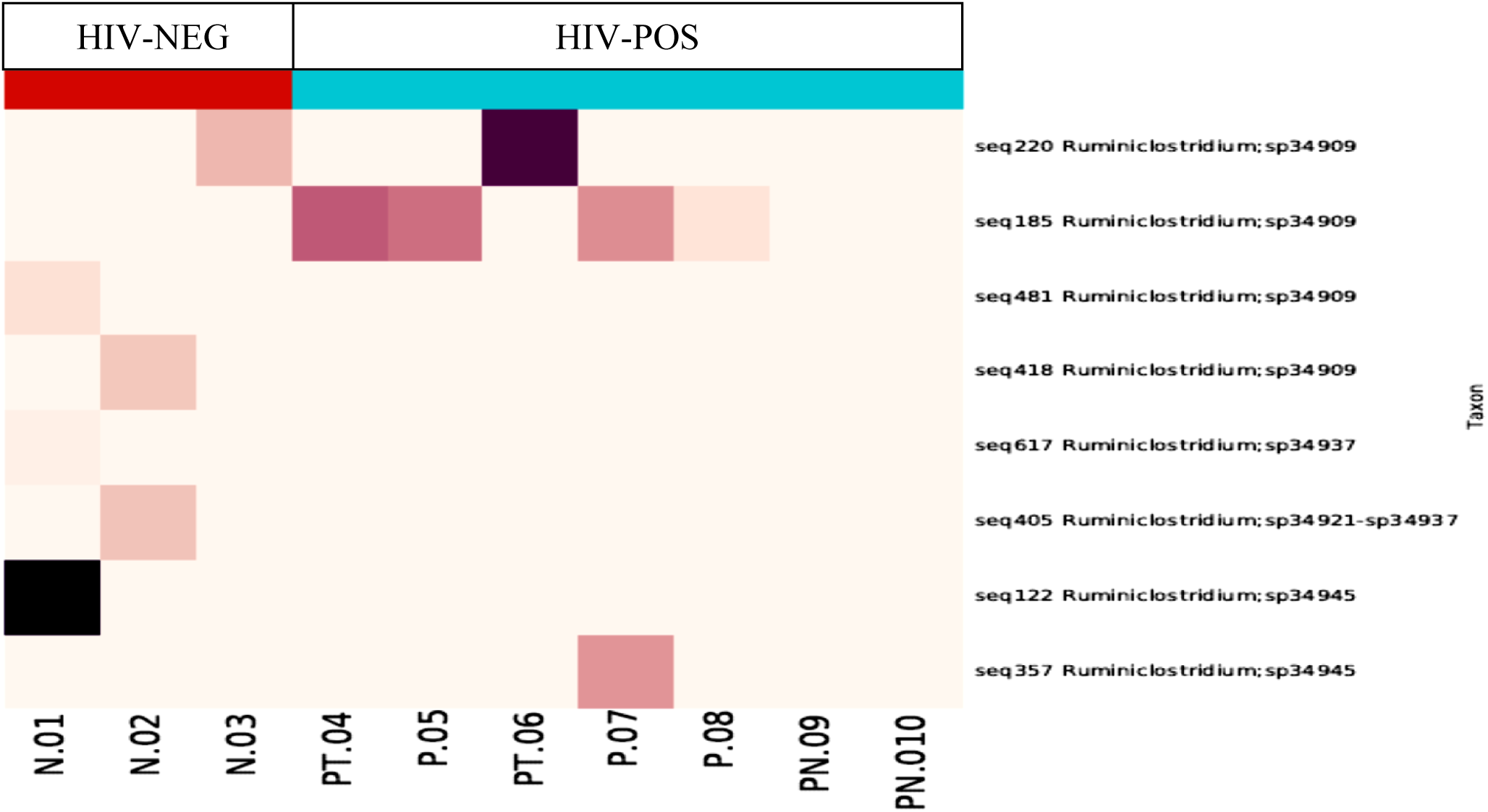
*Ruminiclostridium* unique taxon sequence distribution. Heat map depicting increased variation of unique sequences for Ruminiclostridium seq 185 and seq 220 amongst HIV-positive individuals on HAART and HIV/TB coinfected individuals

### Gut pathobiont composition in the study participants

Findings from our study showed a significantly high proportion of pathobiont among HIV-negative and HIV-positive individuals. Twenty-eight gut microbiota were recorded as pathogenic following 16S rRNA analysis. A Majority of the pathobiont were *Enterobacter spp, Citrobacter spp, Clostridium perfringens, Enterococcus spp, Paraclostridium spp, and Esherichia-Shigella coli*.

The least observed pathogenic strains included *Haemophilus parainfluenzae, Klebsiella spp, Moganella morganii, leuconostoc lactis, Aeromonas sanareli, and Campylobacter consisus* (**Figure 14**). The proportion of gut pathobiont was relatively higher among HIV-negative study participants when compared to their HIV-positive counterparts.

**Figure 14:**
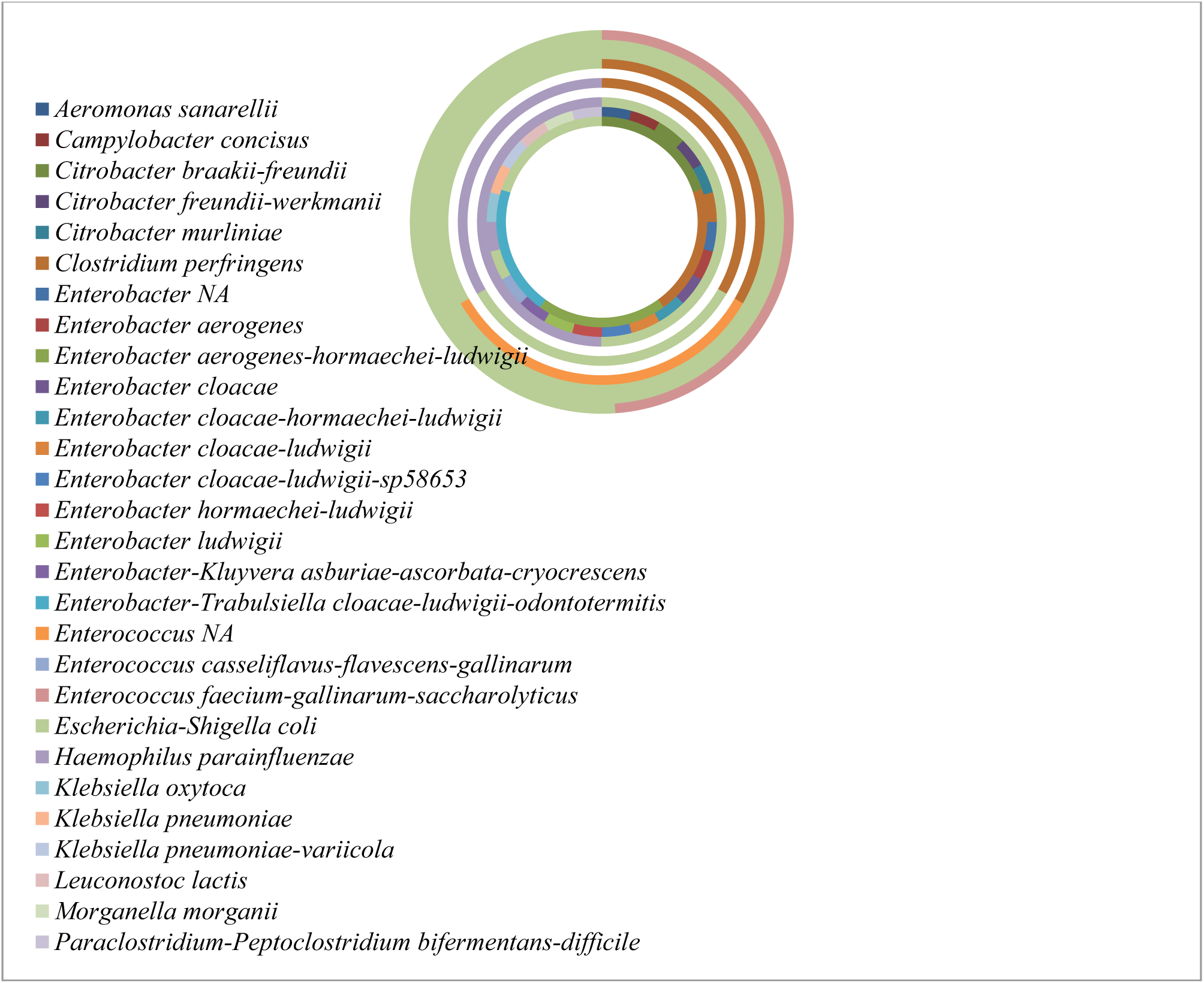
Gut pathobionts amongst the study participants.

## Discussion

We have described a database for the gut microbiome in terms of OTU abundance in a Cameroonian population residing in the South West Region of the country. These findings are the first of their kind, making available a profile of 347 gut microbiome bacterial species present in this community. Particularly, from the 347 gut microbiome bacterial species profiled, eight phyla were identified, with 55 unique species/oligotypes containing more than one sequence and 28 pathobionts. A host of unknown/unclassified gut microbiome bacterial species were also noted circulating among the study population. This study has not only described the type and frequency of gut microbiome bacterial species in the South West Region, but it has also put forward the opinion that the core microbiome might be subjective to geographical settings. Thus, the functionality of the gut microbiome bacterial species in a geographical setting might vary from those of other areas. The majority of the gut microbial community originated from the phylum Firmicutes followed by Bacteroidetes, Proteobacteria, Actinobacteria, Fusobacteria while the least were Euryarchaeota, Synergistetes, and Verrucomicrobia. This observation was similarly reported by Cheng *et al*. [21] who observed that the microbiota profile with the most dominant taxa were Firmicutes, Bacteroidetes, and Actinobacteria in both HIV-infected and uninfected individuals. A high proportion of unclassified gut microbial communities were also recorded amongst the study population.

At the genus level, the gut microbiome communities in HIV-negative individuals were more diverse compared to HIV-positive individuals. This was previously reported by Vázquez-Castellanos *et al*. [22] in which Healthy subjects clustered separately from positive subjects based on their 16S rRNA sequencing. In HIV-positive individuals, higher proportions of four microbial communities: *Parabacteroides distasonis, Prevotella copri, Blautia wexlerae, Lachnoclostridium sp32400*, and *Faecalibacterium prausnitzzi* were recorded from our study. These findings are inconsistent with the work of Lozupone *et al*. [23] who observed a low proportion of *Parabacteroides distasonis*, and an increase in the member of the *Prevotella* family. Contrary to our study, Dillon *et al*. [10] found a significant decrease in the Becteroidaceae and Lachnospiraceae families in HIV-positive individuals versus healthy controls.

Regarding pathobionts, 28 were recorded from this study participants. However majority of the HIV-negative individuals, presented with more pathobionts as compared to HIV –infected individuals. Similar pathobionts were observed in high proportion but included an extended spectrum of other communities in some other studies.. Contrarily to our study, Zhou *et al*., [24] reported higher proportions of potentially pathogenic microbes such as Proteobacteria, *Enterococcus, Streptococcus, Lactobacillus, and Ruminococcus*. Lower levels of *Bacteroides vulgatus, Prevotella copri-sp13942, Megamonas funiformis Anaerostipes hadrus, Coprococcus comes-sp32193, Lachnoclostridium sp32343-sp32393-sp32423, Faecalibacterium sp34558, Subdoligranulum sp35380-sp35585, Prevotella stercorea, Blautia sp32056 Eubacterium hailli, Ruminiclostridium sp34921-sp34937, Bilophila wadsworthia, Escherichia-shigella coli, Senegalimassilia anaerobia, Bacteroides fragilis-ovatus, Bacteroides uniformis, Sutterella wadsworthensis, Holdemanella biformis, Holdemanella sp36738, Subdoligranulum sp35580-sp35585-variabile, Romboutsia ilealis, Roseburia intestinalis, Blautia sp32002, Blautia-lachnoclostridium sp32052-sp32410, Fusicatenibacters saccharivorans, and Lachnospira sp32454* were observed in HIV-positive as compared to HIV-negative individuals in this study. Comprehensively, the latter microbiome occurrence at the taxa level of individual gut microbiota genus was lower with all HIV-positive patients irrespective of HAART and cotrimoxazole treatment, when compared with those of HIV negative individuals, suggesting the direct effect of HIV infection in promoting dysbiosis.

Studies have reported lower counts of *Lactobacillus* [25] and *Bifidobacterium* in the stool of HIV-treatment naïve individuals [26]. Previous works have linked the depletion of *Bifidobacterium* and *Lactobacillus* during HIV infection and their consequential effects on gut barrier destruction and poor immune function in the GALT [27]. The gut microbiota of patients with HIV, compared with those of controls from this study, contained lower levels of *Bacteroidetes, Prevotella, Megamonas, Dialister, Ruminiclostridium, Faecalibacterium, Ruminococcus, Lachnospira, Roseburia, Blautia, Bacteroides vulgatus, Bacteroides uniformis, Phascolartobacterium faeclum, Ruminococcus bromii* and *Bacteroides stercoris*.

Dillon *et al*. [10] observed an increased abundance of the phylum Proteobacteria in HIV-infected individuals. However, in the present study, we did not observe an increased abundance of the phylum Proteobacteria in HIV-infected patients using fecal samples. These findings are inconsistent with previous studies which largely focused on populations from Western countries [10]. This variation in the composition of the gut microbiota might reflect differences in diet or host genetic background between the Cameroonian and Western populations. The Western diet is high in fat and calories, while the diet of individuals in the Southwest Region of Cameroon typically includes relatively low levels of fat, sugar, and meat, which may have significant effects on the gut microbiota. Kashyap *et al*. [28] have suggested that different dietary patterns are strongly associated with gut microbiota enterotypes. Lozupone *et al*. [22] have shown that diets high in fat and protein and low in carbohydrates and fiber are correlated with the loss of beneficial bacteria in HIV-infected patients. This study affirms that fiber diets are correlated with the loss of beneficial bacteria in HIV-infected patients.

Identification and profiling for unique amplicon sequences/oligotype in the sample population were carried out through Taxa2SV analysis and it was the first-ever conducted study in the Southwest Region of Cameroon. There were sequence variations for 55 gut microbiota genera amongst HIV-positive and HIV-negative individuals (*Alloprevotella, Veillonella, Sutterella, Subdoligranulum, Streptococcus, Ruminococcus, Ruminiclostridium, Roseburia, Romboutsia, Prevotella, Phascolarctobacterium, Paraprevotella, Parabacteroides, Megasphaera, Megamonas, Lachnospira, Lachnoclostridium, Klebsiella, Intestinibacter, Holdemanella, Haemophilus, Fusicatenibacter, Faecalibacterium, Eubacterium, Escherichia-Shigella, EnterobacterKluyvera, Enterobacter, Dorea, Coprococcus, Collinsella, Clostridium, Catenibacterium, Butyrivibrio, Blautia, Bifidobacterium, Bacteroides, and Anaerostipes*). In this analysis, a high proportion of Unique amplicon sequences were found in HIV-infected individuals when compared with HIV-negative counterparts. A majority of the taxon in this study contained more than one unique sequence with some protracting far distance genotypic makeup from each other. The increased presence of unique gut microbiome sequence distribution among HIV infected, HIV/TB co-infected and HIV negative individuals in this Cameroonian population warrants further study.The increased occurrences of these unique gut microbiome sequences with HAART, ATB, and cotrimoxazole treatment, might indicate the possible involvement of these drugs in the acquisition of new strains of these gut flora.

In summary, our study affirms the alterations in the gut microbiome and dysbiosis in HIV-infected patients from the Cameroonian Population with the emergence of unique gut microbiome species. In this study, HIV-negative individuals had a more rich microbial community as compared to HIV patients, which had a more diverse population with unknown species and classification. contrary findings have been reported in previous studies, indicating lower α-diversities of intestinal microbiota among HIV-infected individuals [11, 29], while such discrepancies were not statistically significant in some studies [30]. Going forward, we need to disentangle three separate notions: HIV presence, HAART/Cotrimoxazole, and HAART/ATB can to some degree exert pressure on the gut leading to HIV-associated gut dysbiosis as shown in most but not all studies. However this pressure exerted on the gut could lead to the emergence of new microbial species which can also lead to a separate dysbiosis, which may be confounded by the HIV-associated gut dysbiosis and go unreported and unstudied.

## Conclusion

Within the limitations of the study the following conclusions can be made:

Gut microbiome bacterial comprising 347 strains were present amongst study participants in the Cameroonian popolation. Of the 347 strains, 55 unique species containing more than one sequence, 28 pathobionts, and a few unknown/unclassified gut microbiome bacterial species were observed. The phylum of Gut microbiota communities identified included Firmicutes, Bacteroidetes, Proteobacteria, Actinobacteria, Fusobacteria, Euryarchaeota, Synergistetes, Verrucomicrobia, and unclassified phylum. The gut microbiome OTU diversity for HIV-negative individuals was abundant with less enrichment of butyrate-producing flora as compared to those of HIV-positive, which was less diverse with increased enrichment of butyrate-producing flora. Also the unique gut microbiome/oligotype OTU was significantly higher amongst HIV-positive individuals on HAART, ATB, and cotrimoxazole as compared to HIV-positive treatment naïve individuals. Further works on functionality of the gut microbiome, the origin of pathobionts and cause of the absence and or the emergence of new strains of unique gut microorganisms needs to be investigated.

## Data Availability

The raw sequencing data was zipped and can be accessed at: https:/epiquest.s3.amazonaws.com/epiquest zr2768/CMCDFQUJAMKPZKAFWSY3FJLBJDRTBYVE/rawdata/zr2768.rawdata.190904.zip

https:/epiquest.s3.amazonaws.com/epiquestzr2768/CMCDFQUJAMKPZKAFWSY3FJLBJDRTBYVE/rawdata/zr2768.rawdata.190904.zip

## Funding source

This research did not receive any specific grant from funding agencies in the public, commercial, or not-for-profit sectors.

## Declaration of Competing Interest

The authors have no conflict of interest.

## Acknowledgment

We are thankful to the Department of Medical Laboratory Science, University of Buea (Cameroon), for providing an instrumental facility at the Medical Research and Bacteriology Laboratory (FHS-MRBL) and the Infectious Disease Laboratory, Faculty of Health Sciences, University of Buea for DNA extraction. We are also grateful to the Buea Regional Hospital and UPEC unit for their facilities and services provided. We also acknowledge the ZYMO Research Cooperation USA (Zymo Research, Irvine, CA), for their services rendered in the Library Preparation (16S V3-V4), Pooling, and Post-Library QC, and Illumina MiSeq® Sequencing(2×300) and also for Bioinformatics analysis.

## Author Contribution Statement

SEA, EAA, CNN, TBP, and JNA conceived and designed the research. SEA, CN, EJE, CFN, and MGM conducted experiments. SEA contributed new reagents or analytical tools. SEA, TBP, MGM, WGF, and NMY wrote the manuscript. All authors read and approved the manuscript.

## Notes

### Competing Interest Statement

The authors have declared no competing interest.

### Funding Statement

NO EXTERNAL FUNDING WAS ACQUIRED FOR THIS STUDY. THE PROJECT WAS SPONSORED BY THE CORRESPONDING AUTHOR

### Author Declarations

Institutional Review Board (IRB) of the Faculty of Health Sciences (FHS) of the University of Buea, Cameroon Ref N°: 2018/826-06/UB/SG/IRB/FHS

## REFERENCES

1. Turnbaugh PJ, Ley RE, Hamady M, et al. The human microbiome project. Nature, 2007;449:804–810.

2. Jay L, Brett W, Daniel F, Stephanie M D, Cara C W, and Alan LL. Inside Out: HIV, the Gut Microbiome, and the Mucosal Immune System. J Immunol 2017; 198:605–614.

3. Gill, S.R., Pop, M., DeBoy, R.T., Eckburg, P.B., Turnbaugh, P.J., Samuel, B.S. et al. Metagenomic analysis of the human distal gut microbiome. Science, 2006; 312, 1355–1359

4. Sender, R., Fuchs, S. and Milo, R. Revised estimates for the number of human and bacteria cells in the body. PloS Bio. 2016;14(8):e1002533.

5. Jiang, H., Ling, Z., Zhang, Y., Mao, H., Ma, Z., Yin, Y. et al. Altered fecal microbiota composition in patients with major depressive disorder. Brain Behavior and Immunity, 2015;48: 186–194

6. Simon Eyongabane AKO, Eric Achidi AKUM, Céline Nguefeu NKENFOU, Jules Clement N. ASSOB, Thumamo Benjamin POKAM. In-Vitro Susceptibility of Gut Pathobiont associated with Microbial Translocation to Cotrimoxazole and Antiretroviral. Scientific African, 6 (2019) e00192

7. Cameroon/UNAIDS 2018. [Internet] Available at https://www.unaids.org/en/regionscountries/cameroon

8. Marcel Tongo, Darren P Martin, Lycias Zembe, Eitel Mpoudi-Ngole, Carolyn Williamson, Wendy A Burgers. Characterization of HIV-1 gag and nef in Cameroon: further evidence of extreme diversity at the origin of the HIV-1 group M epidemic Virol J. 2013; 10: 29. Published online 2013 Jan 22. DOI: 10.1186/1743-422X-10-29 PMCID: PMC3560183

9. Chow J., Tang H and. Mazmanian S. K. Pathobionts of the gastrointestinal microbiota and inflammatory disease. Curr. Opin. Immunol., 2011;23: 473–480.

10. Dillon, S. M., E. J. Lee, C. V. Kotter, G. L. Austin, Z. Dong, D. K. Hecht, S. Gianella, B. Siewe, D. M. Smith, A. L. Landay, et al. An altered intestinal mucosal microbiome in HIV-1 infection is associated with mucosal and systemic immune activation and endotoxemia. Mucosal Immunol. 2014; 7: 983–994

11. Mutlu, E. A., A. Keshavarzian, J. Losurdo, G. Swanson, B. Siewe, C. Forsyth, A. French, P. Demarais, Y. Sun, L. Koenig, et al. A compositional look at the human gastrointestinal microbiome and immune activation parameters in HIV infected subjects. PLoS Pathog., 2014; 10: e1003829

12. Simon Eyongabane Ako, Eric Achidi Akum, Jules Clement N. Assob, Céline Nguefeu Nkenfou, Pokam Thumamo Benjamin, Enoh Jude Eteneneng,Cho Frederick Nchang and Mbanya Gladice Mbanya Gut Microbiota Dysbiotic Pattern and Its Associated Factors in a Cameroonian Cohort with and without HIV Infection. Journal of Advances in Microbiology, 17(1): 1–23, 2019; Article no.JAMB.49827

13. Respess RA, Rayfield MA and Dondero TJ (2001) Laboratory testing and rapid HIV assays applications for HIV surveillance in hard-to-reach populations. AIDS 15 Supplement 3: S49–S59.

14. Julius Y. F, Alfred K. N, Charles K, Fang Q, Dora M., et al. Adherence to Antiretroviral Therapy (ART) in Yaoundé-Cameroon: Association with Opportunistic Infections, Depression, ART Regimen and Side Effects. PLoS One. 2017; 12(1): e0170893

15. Neogi, U., Gupta, S., Rodridges, R., Sahoo, P. N., Rao, S. D., Rewari, B. B., Shet, A.. Dried blood spot HIV-1 RNA quantification: a useful tool for viral load monitoring among HIV-infected individuals in India. The Indian journal of medical research (2012), 136(6), 956–962.

16. Shantelle C, Elloise du T, Mamadou K, Clinton M, Heather J. Z, Mark P. N. A comparison of the efficiency of five different commercial DNA extraction kits for extraction of DNA from faecal samples. J Microbiol Methods. 2013 Aug; 94(2): 103–110. doi: 10.1016/j.mimet.2013.05.008

17. Callahan B.J., McMurdie P.J., Rosen M.J., Han A.W., Johnson A.J., Holmes S.P., (2016) DADA2: High resolution sample inference from Illumina amplicon data. Nat Methods 13(7):581–3.

18. Caporaso, J.G., Kuczynski, J., Stombaugh, J., Bittinger, K., Bushman, F.D., Costello, E.K. et al. (2010) QIIME allows analysis of high-throughput community sequencing data. Nat Methods 7: 335–336.

19. Segata, N., Izard, J., Waldron, L., Gevers, D., Miropolsky, L., Garrett, W.S., and Huttenhower, C. Metagenomic biomarker discovery and explanation. Genome Biol. 2011; 12: R60.

20. Leandro NL, Roberta R F, Eric WT, Luiz FW, Rethinking microbial diversity analysis in the high throughput sequencing era. Journal of Microbiological Methods. 2011;86:1 Pages 42-51

21. Soo C L, Ling L C, Siew H Y, Tsung F K, Chan Y L, Raja I R A et al. Enrichment of gut-derived Fusobacterium is associated with suboptimal immune recovery in HIV-infected individuals. Scientific reports, 2018; 8:14277

22. Vázquez-Castellanos JF, Serrano-Villar S., Latorre A., Artacho A., Ferrus ML.,Madrid N et al. Altered metabolism of gut microbiota contributes to chronic immune activation in HIV-infected individuals. Mucosal immunol., 2015;8(4);760–72

23. Lozupone CA, Matthew ER, Neff CP, Fontenot AP, Thomas BC & Palmer BE. HIV-induced alteration in gut microbiota, Gut Microbes. 2014; 5:4, 562-570,

24. Zhou Y., Zech Z., Chen Y et al. Gut microbiota offers universal biomarkers across ethnicity in inflammatory bowel disease diagnosis and infliximab response production. mSystems, 2018;3(1):e00188–17

25. González-Hernández LA, Jave-Suarez LF, Fafutis-Morris M, et al. Synbiotic therapy decreases microbial translocation and inflammation and improves immunological status in HIV-infected patients: a double-blind randomized controlled pilot trial. Nutr J. 2012;11:90.

26. Luz A. González-Hernández, Mariana del Rocio Ruiz-Briseño, Karina Sánchez-Reyes, Monserrat Alvarez-Zavala, Natali Vega-Magaña, Alvaro López-Iñiguez1, Julio A. Díaz-Ramos et al. Alterations in bacterial communities, SCFA and biomarkers in an elderly HIV-positive and HIV-negative population in western Mexico. BMC Infectious Diseases. 2019; 19:234.

27. Nwosu FC, Avershina E, Wilson R, Rudi K. Gut microbiota in HIV infection: implication for disease progression and management. Gastroenterol Res Pract. 2014;803185.

28. Kashyap PC, Marcobal A, Ursell LK, et al. Genetically dictated change in host mucus carbohydrate landscape exerts a diet-dependent effect on the gut microbiota. Proc Natl Acad Sci USA. 2013; 110: 17059–64.

29. Nowak, P., M. Troseid, E. Avershina, B. Barqasho, U. Neogi, K. Holm, J. R. Hov, K. Noyan, J. Vesterbacka, J. Svärd, et al. Gut microbiota diversity predicts immune status in HIV-1 infection. AIDS 2015; 29: 2409–2418.

30. Li, S. X. Armstrong A, Neff CP, Shaffer M, Lozupone CA, Palmer BE. Complexities of gut microbiome dysbiosis in the context of HIV infection and antiretroviral therapy. Clin. Pharmacol. Ther. 2016;.99: 600–611

